# Post-Exposure Prophylaxis with SA58 (anti-COVID-19 monoclonal antibody) Nasal Spray for the prevention of symptomatic Coronavirus Disease 2019 in healthy adult workers: A randomized, single-blind, placebo-controlled clinical study

**DOI:** 10.1101/2022.12.28.22283666

**Authors:** Rui Song, Gang Zeng, Jianxing Yu, Xing Meng, Xiaoyou Chen, Jing Li, Xiaoliang Xie, Xiaojuan Lian, Zhiyun Zhang, Yunlong Cao, Weidong Yin, Ronghua Jin

## Abstract

**BACKGROUND:** This study has assessed a new Anti-COVID-19 Monoclonal Antibody Nasal Spray (SA58) for post-exposure prophylaxis (PEP) against symptomatic coronavirus disease 2019 (COVID-19).

**METHODS:** We conducted an efficacy study in adults aged 18 years and older within three days of exposure to a SARS-CoV-2 infected individual. Recruited participants were randomized in a ratio of 3:1 to receive SA58 or placebo. Primary endpoints were laboratory-confirmed symptomatic COVID-19 within study period.

**FINDINGS:** A total of 1,222 participants were randomized and dosed (SA58, n=901; placebo, n=321). Median of follow-up was 2·25 days and 2·79 days for SA58 and placebo, respectively. Adverse events occurred in 221 of 901 (25%) and 72 of 321 (22%) participants with SA58 and placebo, respectively, with no significant difference (P=0·49). All adverse events were mild in severity. Laboratory-confirmed symptomatic COVID-19 developed in 7 of 824 participants (0·22 per 100 person-days) in the SA58 group vs 14 of 299 (1·17 per 100 person-days) in the placebo group, resulting in an estimated efficacy of 80 · 82% (95%CI 52 · 41%-92 · 27%). There were 32 SARS-CoV-2 RT-PCR positives (1·04 per 100 person-days) in the SA58 group vs 32 (2·80 per 100 person-days) in the placebo group, resulting in an estimated efficacy of 61·83% (95%CI 37· 50%-76· 69%). A total of 21 RT-PCR positive samples were sequenced. 21 lineages of SARS-CoV-2 variants were identified, and all were the Omicron variant BF·7.

**INTERPRETATION:** SA58 Nasal Spray showed favorable efficacy and safety in preventing SARS-CoV-2 infection or symptomatic COVID-19 in healthy adult workers who had exposure to SARS-CoV-2 within 72 hours.

**FUNDING:** No funding was received for this study.

**Research in context:** *Evidence before this study:* Monoclonal antibodies (mAbs) and the post-exposure prophylaxis (PEP) with mAbs represent a very important public health strategy against COVID-19 outbreak. SA58 Nasal Spray is a broad-spectrum anti-COVID-19 mAb, developed by Sinovac Life Sciences Ltd. for treatment and prophylaxis against COVID-19. SA58 has been shown to potently neutralize ACE2-utilizing sarbecoviruses, including most of circulating Omicron variants. We searched PubMed on Nov 21, 2022, for published clinical trials, with no language or date restrictions, using various combinations of the search terms of “monoclonal antibodies”, “SARS-CoV-2”, “COVID-19”, “prophylaxis”, and “prevention”. Three published trials were identified. The first study reported the efficacy of AZD7442 (tixagevimab/cilgavimab) PEP against symptomatic COVID-19 in adults aged ≥18 years over a 183-day follow-up period. The primary efficacy end point of post-exposure prevention of symptomatic COVID-19 was not met, though AZD7442 showed promising results in participants who were SARS-CoV-2 RT-PCR negative at baseline. The second study reported the efficacy and safety of bamlanivimab for COVID-19 prevention in household contacts of individuals with a SARS-CoV-2 infection in a high-risk transmission setting over a one-month efficacy assessment period. The third study reported REGEN-COV (casirivimab/imdevimab) for preventing symptomatic Covid-19 and asymptomatic SARS-CoV-2 infection in previously uninfected household contacts of infected persons. Both bamlanivimab and REGEN-COV showed satisfactory safety profile and efficacy against COVID-19 and were licensed for PEP use in the U.S. However, due to the circulating Omicron variants have developed significant escape properties, the emergency use authorization of bamlanivimab and REGEN-COV for treatment and PEP against COVID-19 has been discontinued by the U.S. Food and Drug Administration. Till the end of December 2022, no drug was available for PEP use against COVID-19.

*Added value of this study:* During a recent large outbreak of the Omicron BF·7 sublineage in Beijing, our preliminary results in healthy adults within 72 hours of contact with SARS-CoV-2-infected individuals showed that SA58 nasal spray was highly effective in preventing symptomatic COVID-19 and SARS-CoV-2 infection caused by the sublineage, which variants have shown significant escape of immunity in previous studies. SA58 was able to significantly lower the risk of laboratory-confirmed COVID-19 by 80·82% (95%CI 52·41%-92·27%) and of SARS-CoV-2 infection by 61·83% (95%CI 37·50%-76·69%) in our study participants.

*Implications of all the available evidence:* This trial showed the ability of a nasal spray of broad-spectrum anti-COVID-19 mAb SA58 to provide satisfactory protection against one circulating Omicron strain of SARS-CoV-2. The drug had a favorable safety profile and was well tolerated by healthy adults. This newly developed mAb is resistant to SARS-CoV-2 mutations and may provide a new powerful countermeasure to tackle against the immunity-escaping variants of SARS-CoV-2 circulating in the population. The intranasal administration of SA58 is novel and has many advantages over intramuscular injections of mAbs previously licensed, as it is less invasive and more acceptable in recipients. Auto-administration with easiness of use may allow early administration, probably a key feature for prevention.

## Introduction

Postexposure prophylaxis (PEP) is the administration of chemicals or immunotherapeutic agents to prevent the development of infection or to slow the illness progression prior to the illness onset. PEP has been routinely recommended for several viral infections, including influenza virus,^1^ rabies virus,^2^ human immunodeficiency virus,^3^ hepatitis B virus^4^ and varicella-zoster virus,^5^ especially for those who have higher risks of severe outcomes or mortality following infection.^6^

Coronavirus disease 2019 (COVID-19) is a contagious condition caused by severe acute respiratory syndrome coronavirus 2 (SARS-CoV-2) that firstly emerged in December 2019.^7^ The on-going COVID-19 pandemic has led to high morbidity and mortality worldwide,^8^ resulting in 648 million laboratory-confirmed cases and 6· 64 million deaths globally as of December 15, 2022.^9^ This substantial impact of COVID-19 has reshaped the world and profoundly changed public health practices, including the development of various preventions (e.g., vaccines) and therapeutic measures. Several passive immunotherapeutic antibodies against SARS-CoV-2 have since been generated, tested, and moved into clinical trials.^10^ However, due to the high frequency of mutations, newly emerged SARS-CoV-2 variants have been circulating in the population (e.g., Omicron variants). These variants have developed significant escape properties, resulting in several monoclonal antibodies (mAbs) that had initially been authorized to treat COVID-19 or use as PEP, to be discontinued.^11^ Till the end of December 2022, Evusheld (tixagevimab/cilgavimab) was the only mAb licensed by FDA for emergency use as pre-exposure prophylaxis for prevention of COVID-19 in adults and pediatric individuals (12 years of age and older weighing at least 40 kg).^12^ Although a phase 3 trial assessing Evusheld (tixagevimab/cilgavimab) for PEP against symptomatic COVID-19 showed promising results,^13^ no drug have since been approved as PEP use against COVID-19. As broad-spectrum mAbs and PEP represent a very important public health strategy against COVID-19 outbreak, especially among high-risk populations who are vulnerable to severe disease following infection, it is important to develop and evaluate more potential broad-spectrum mAbs, which can be protective against the SARS-CoV-2 Omicron sub-lineages circulating in the population and other upcoming variants.

SA58 Nasal Spray, a broad-spectrum anti-COVID-19 mAb, was developed by Sinovac Life Sciences Co., Ltd. This antibody was identified from a large collection of broad sarbecovirus mAbs isolated from SARS-CoV-2-vaccinated SARS convalescents. It has been shown to potently neutralize ACE2-utilizing sarbecoviruses, including circulating Omicron variants (BA·1, BA·2, BA·2· 12·1, BA·3, BA·4/BA·5, BF·7, and other variants that tested so far) in in vitro neutralizing and in animal challenge studies.^14^ The early pharmacokinetic results in human volunteers showed that SA58 was safe, and that the half-life of SA58 administered intranasally was 2-4 hours in the nasal cavity and 12-27 hours in the nasopharynx, with no detectable drug components in the blood (below the detection limit of the method used). In previous study on the high-risk population of medical workers, the most common encountered symptoms were runny nose, nasal mucosal dryness, nasal congestion, and headache post administration, suggesting good safety and tolerability of a SA58 Nasal Spray. More information about the effect of SA58 Nasal Spray administered as PEP against symptomatic COVID-19 or SARS-CoV-2 infection was needed. Therefore, the aim of this study was to estimate the efficacy of SA58 nasal spray in preventing symptomatic COVID-19 in healthy adults who had exposure to individuals with laboratory-confirmed SARS-CoV-2 infection within 72 hours.

## METHODS

### Study Design and Participants

This randomized, single-blind, placebo-controlled clinical trial evaluated the efficacy and safety of the SA58 nasal spray in healthy adult workers within 72 hours of contact with SARS-CoV-2-infected individual in Beijing, China. The study was conducted from November 26^th^, 2022 to December 9^th^, 2022 at 21 construction sites (median number of workers=45, range= 9-235) that had COVID-19 outbreaks reported within two days of the first COVID-19 case notified. All participants in this study were voluntary and provided written informed consent before enrolment. The clinical trial protocol and informed consent form were approved by the Ethics Committee of Beijing Ditan Hospital, Capital Medical University (reference no., DTEC-YW2022-024-01). The study was registered with ClinicalTrials· gov (NCT05667714).

In this study, all healthy workers in the 21 construction sites were offered the opportunity to participate based on the following inclusion/exclusion criteria. Participants were eligible for inclusion if they were aged 18 years or older, had potential exposure to a well identified individual with laboratory-confirmed SARS-CoV-2 infection (index case), and if the presumable contact occurred within 72 hours of the positive RT-PCR test of the index case. The key exclusion criteria were individuals with known history of severe allergies or reaction to any component of inhaled SA58 nasal spray; those currently pregnant, lactating, or expected to be pregnant during the study period; those who participated in any kind of clinical trials of SARS-CoV-2 neutralizing antibody injections in the preceding 180 days before screening or participated in any investigational medicinal product in the preceding four weeks before screening; were unable to take nasal spray inhalation; had reported fever at enrolment or axillary temperature of more than 37·0 °C; had severe neurological disease (e.g., epilepsy, convulsions, or seizures) or psychosis, or family history of psychosis; or had any other significant chronic disease, disorder, or finding that, in the judgment of the investigator, significantly increased the risk to the participant because of participation in the study, affected the ability of the participant to participate in the study, or impaired interpretation of the study data. Full eligibility criteria are provided in the Supplementary Material. A nasopharyngeal swab, nasal swab, or throat swab was collected at baseline for detection of SARS-CoV-2 nucleic acid by RT-PCR tests, but the results of baseline RT-PCR tests were not used to determine the eligibility of participants.

### Outcomes

The primary efficacy endpoint was the laboratory-confirmed symptomatic COVID-19 that occurred during the case monitoring period between 24h after the first administration and 24h after the last administration of SA58 or placebo. COVID-19 case was defined based on symptoms (see Supplementary Table 1). Severe COVID-19 was defined based on the Protocol for Prevention and Control of COVID-19 (9th edition) issued by the National Health Commission of the People’s Republic of China, and we arbitrarily defined severe COVID-19 as severe case and very-severe case combined (see Supplementary Table 2).^15^ The safety endpoints included incidence of adverse events (AEs), serious AEs (SAEs), and AEs of special interest (AESIs).

### The investigational drug

The investigational drug SA58 was prepared into 2 ml prefilled sprayer (20 sprays per bottle), containing 5 mg of anti-COVID-19 mAb per ml. Placebo, only without anti-COVID-19 mAb as compared with SA58 nasal spray, was also prefilled into bottles with identical package that could not be easily distinguished by their appearance. The drug and placebo were self-administered by nasal sprays with a video instruction. When used, insert the sprayer nozzle into each nose nostril and press the pump to spray 0· 1 ml of the nebulized solution into the nasal cavity. Each administration of the drug consisted of two sprays with one spray in each nostril, and a total of 1 mg antibody was administered into both nostrils. For an ordinary exposure day, 5-6 administrations of SA58 or placebo were recommended at an interval of 3-4 hours per administration, with the last administration given before going to bed.

### Procedures

#### Randomization and Masking

After enrolment, participants were randomized in a ratio of 3:1 to receive SA58 or placebo at the study site. A statistician (independent of the study) assigned treatment at random using a standard computer pseudorandom number generator. Only the participant and the organizer of the site were blinded and unaware of what had been allocated.

#### Administration of drugs

After confirmation of participants’ contact to a laboratory-confirmed SARS-CoV-2 infection, participants were classified into two groups according to their type of exposure to SARS-CoV-2, e.g., Group A participants with continuous exposure to COVID-19 in which potential contact to SARS-CoV-2 infected individual was not blocked by removing or eliminating the source of infection in the site, and Group B participants with one-time exposure in which contact to SARS-CoV-2 infected individual was immediately blocked by managing study participants in isolation facilities for highly infectious Diseases.

For the purpose of PEP against continuous exposure to COVID-19, Group A participants were administered SA58 or placebo on every exposure day according to the recommended administration schedule and stopped administration three days after the source of infection had been eliminated or removed from the study site (e.g., all SARS-CoV-2 infected individual in the site had no symptoms and had negative RT-PCR testing results of SARS-CoV-2 for the last two consecutive days); for the purpose of PEP against one-time exposure in which the contact to the SARS-CoV-2 infected individual was immediately blocked, each participant was administered SA58 or placebo for a maximum of three days according to the recommended administration schedule.

#### Follow-up of Adverse Events

Participants self-monitored for safety post administration and spontaneously reported adverse events (e.g., runny nose, sneezing, nasal congestion, nasal dryness, fever, headache, and fatigue) post administration to study follow-up personnel. Predefined symptoms (solicited events) and other unspecified symptoms (unsolicited events) reported by the participants during the study period were recorded and verified at regular visits by the study investigators. Any SAEs and AESIs (e.g., allergic reaction, autoimmune reaction, and nasal and throat AEs of Grade two and above) were reported up to 30 days since enrolment. Adverse events were graded according to the U.S. National Cancer Institute’s Common Terminology Criteria for Adverse Events (version 5·0).^16^

#### Monitoring of COVID-19 Cases

To specifically measure the incidence of laboratory-confirmed symptomatic COVID-19 post administration, study sites contacted participants daily to collect nasopharyngeal/throat/nasal swab and monitor information on symptoms of COVID-19. The case monitoring period extended from 24 hours after the first administration to 24 hours after the last administration of SA58 or placebo, during which participants were observed closely. If a participant had positive RT-PCR testing result, he/she was defined a SARS-CoV-2 RT-PCR positive participant (e.g., laboratory-confirmed SARS-CoV-2 infection). And if a SARS-CoV-2 RT-PCR positive participant had symptoms of COVID-19 (Supplementary Table 1), this was defined as a laboratory-confirmed symptomatic COVID-19 case. Both laboratory-confirmed SARS-CoV-2 infections and symptomatic COVID-19 cases were followed up to disease resolution, or up to negative RT-PCR tests on two consecutive days.

#### Laboratory methods

The nasopharyngeal/throat/nasal swabs were transferred at 2-8 °C to the central laboratory and tested within 24h of collection. We used real-time reverse transcriptase polymerase chain reaction (RT-PCR) for detecting of SARS-CoV-2 in nasopharyngeal/throat/nasal swabs as per national guidelines.^15^ The RT-PCR positive samples that had cycle threshold values <30 were sequenced to identify SARS-CoV-2 variants by using the methods provided in the Supplementary Material.

### Statistical Analysis

We assumed a 5% secondary attack rate of symptomatic COVID-19 cases in the placebo group during our study period, and a true efficacy of 70% (equating to an attack rate of 1·5% with SA58). Allowing for a dropout rate of 10%, we calculated that a study population of approximately 2300 participants randomized in a 3:1 ratio to treatment or placebo would be sufficient to provide approximately 80% power to demonstrate the lower bound of the two-sided 95% confidence interval (CI) for efficacy to be >0·3.

The incidence rate of symptomatic COVID-19 per 100 person-days were calculated for the SA58 and placebo groups during case monitoring period, by dividing the number of events with the total number of follow-up days. Crude incidence rate ratios (IRRs) comparing the SA58 and placebo groups were calculated. For the adjusted analysis, a Poisson regression model with robust variance using log of follow-up days as an offset to estimate IRR of symptomatic COVID-19 with SA58 vs placebo. Efficacy of treatment was calculated as 1 minus the adjusted IRR and was presented with a two-sided 95% confidence interval (95%CI). In the final efficacy analysis. we excluded participants who tested positive for SARS-CoV-2 by RT-PCR at the screening visit on Day one. Kaplan–Meier curves were also presented for SA58 and placebo groups, with hazard ratios (HR) calculated by using Cox proportional hazard models. We also calculated the efficacy of SA58 against other outcomes, e.g., SARS-CoV-2 infection (SARS-CoV-2 RT-PCR positive during follow-up) and severe COVID-19 following the same methodology as for the primary efficacy end point. For the comparison of individual-level variables at baseline, we used Student’s t-test or Wilcoxon test for continuous variables and the chi-square test or Fisher’s exact test for categorical variables as appropriate. Adverse events were summarized descriptively as frequencies and percentages by type of event and severity. A two-sided P-value of <0· 05 was considered statistically significant. We conducted all analyses in SAS (version 9·1·3).

## Results

### Characteristics of participants

Because of the reopening of China and the lifting of the COVID-19 restrictions, the study participants left study sites and the study ended early before our target sample size was met. In total, 1,694 participants at 21 construction sites were screened and were confirmed to contact with a SARS-CoV-2 infected individual within 72 hours of RT-PCR positive results of an index case. Of which, 60 were ineligible or withdrew early from study before study drug administration, and 1,634 were randomized in a ratio of 3:1 to receive SA58 or placebo. After excluding 401 participants not administrated and 11 participants lost to follow-up, 1,222 participants entered the full analysis set, with 901 participants in SA58 group and 321 in placebo group (Figure 1).

**Figure 1.**
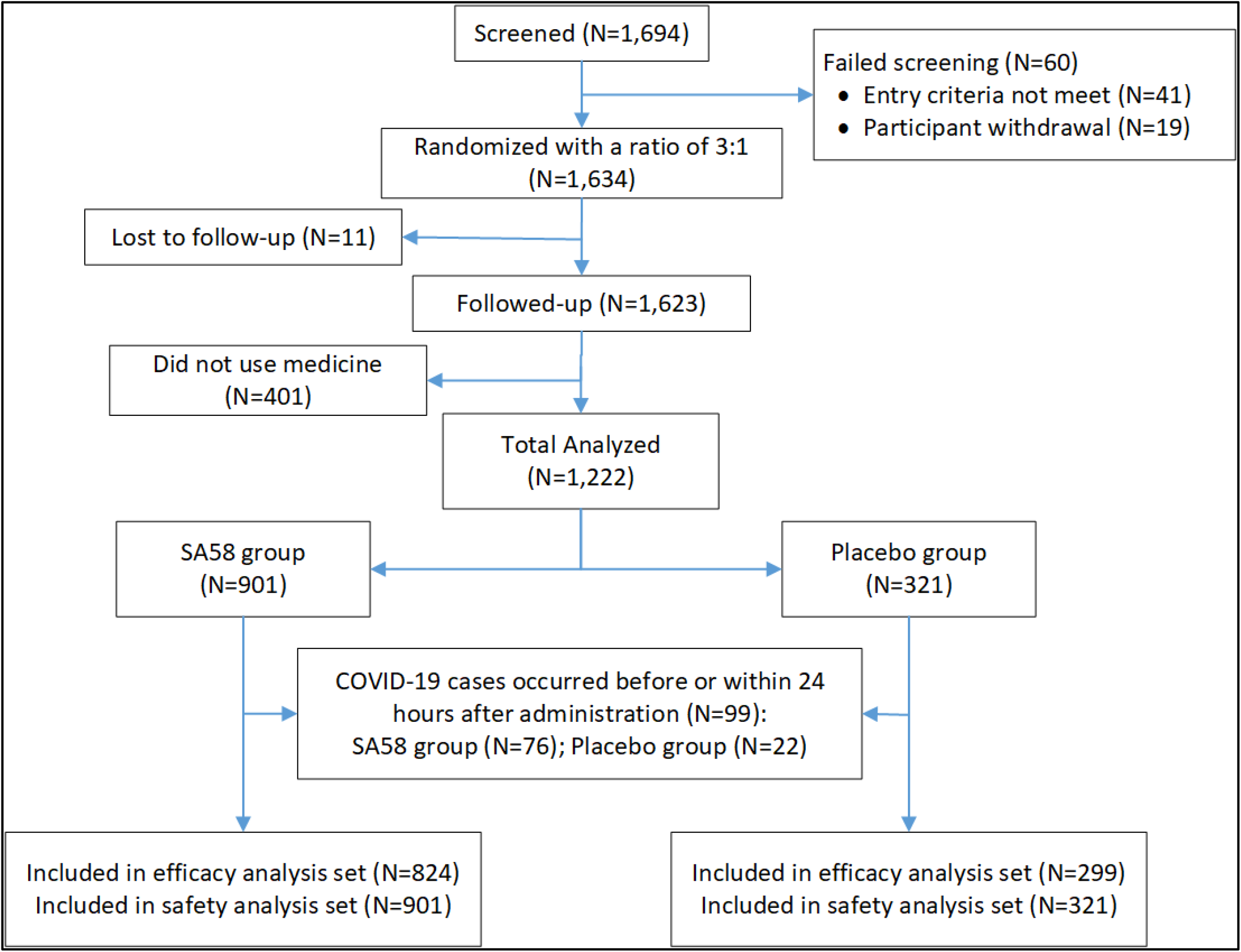
Participant disposition.

In the full analysis set, the median age was 46 years (SD=17 years) in SA58 group and 46 years (SD=17 years) in placebo group, p=0·9934. The majority of participants were male (n=1090, 89%) and adult aged 18-59 years (n=1197, 98%). At baseline, 824 (91%) of 901 participants in SA58 group and 299 (93%) of 321 participants in placebo group were SARS-CoV-2 RT-PCR negative, and no significant difference of SARS-CoV-2 RT-PCR positivity was identified in the two comparison groups. 99 (8%) SARS-CoV-2 RT-PCR positive participants (77 in SA58 and 22 in placebo group) were excluded from the efficacy analysis. Median duration of follow-up for participants was 2·25 days (SD=4·63 days) and 2·79 days (SD=4·67 days) for SA58 and placebo recipients respectively, p=0·1869 (Table 1).

**Table 1.**
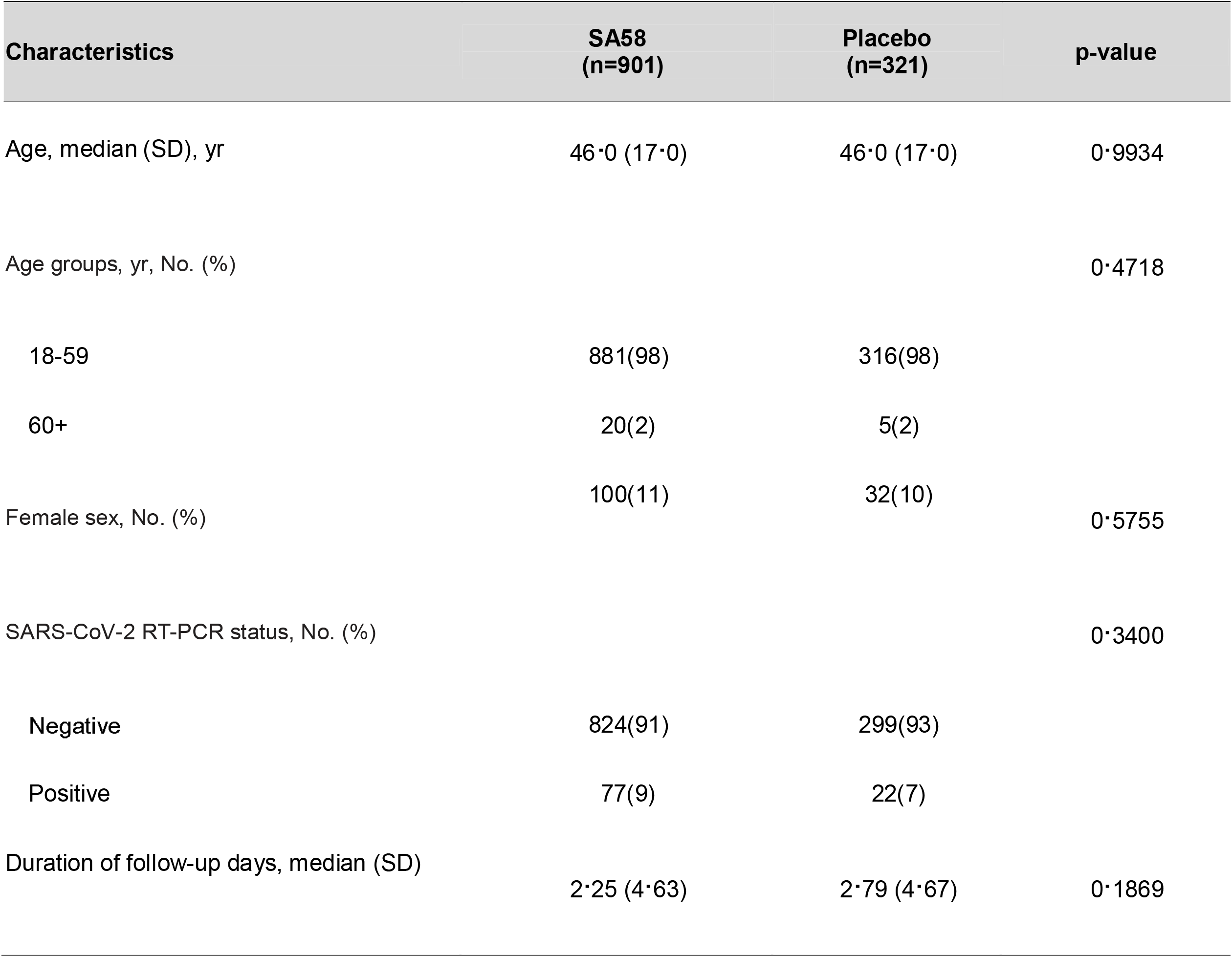
Baseline Characteristics of Study Participants (Full Analysis Set)

### Treatment Efficacy

In the efficacy analysis, 1,123 participants were followed for a total of 4,362 days. The primary outcome laboratory-confirmed symptomatic COVID-19 developed in seven of 824 participants (incidence rate of 0·22 per 100 person-days) in the SA58-treated participants vs 14 of 299 (incidence rate of 1· 17 per 100 person-days) in the placebo group. The occurrence rate of symptomatic COVID-19 was significantly lower for SA58-treated participants vs placebo, with a crude IRR of 0·19 (95%CI 0·08-0·48) and an estimated treatment efficacy of 80· 82% (95%CI 52· 41%-92·27%) (Table 2). Time to first post-administration symptomatic COVID-19 was shown in Figure 2A. The majority of COVID-19 cases developed their symptoms within the first 5 days after enrolment (the median incubation period for COVID-19). The protective effectiveness of SA58 against SARS-CoV-2 infection was also explored by comparing the incidence of SARS-CoV-2 RT-PCR positive between SA58-treated participants vs placebo. There were 32 SARS-CoV-2 RT-PCR positives (incidence rate of 1·04 per 100 person-days) in the SA58-treated participants vs 32 (incidence rate of 2·80 per 100 person-days) in the placebo group, resulting in a crude IRR of 0·38 (95%CI 0·23-0·62) and an estimated SARS-CoV-2 infection lowering efficacy of SA58 of 61·83% (95%CI 37·50%-76·69%) (Table 2). The same occurrence time and pattern of SARS-CoV-2 RT-PCR positivity was observed for SA58-and placebo-treated participants as that of symptomatic COVID-19 cases (Figure 2B). A total of 21 RT-PCR positive samples were sequenced. 21 lineages of SARS-CoV-2 variants were identified, and all were the Omicron variant BF·7 (Supplementary Figure 1).

**Table 2.** Occurrence of SARS-CoV-2 RT-PCR positive and laboratory-confirmed symptomatic COVID-19 in SA58- and placebo-treated participants.

| Variable | SA58 (n=824) |  |  | Placebo (n=299) |  |  | Protective efficacy of treatment (95% CI) |
| --- | --- | --- | --- | --- | --- | --- | --- |
|  | No. of events | Person-days at risk | Incidence rate (per 100 person-days) | No. of events | Person-days at risk | Incidence rate (per 100 person-days) |  |
| SARS-CoV-2 RT-PCR positive | 32 | 3078 | 1·04 | 32 | 1144 | 2·80 | 61·83% (37·50%-76·69%) |
| Symptomatic COVID-19 | 7 | 3162 | 0·22 | 14 | 1200 | 1·17 | 80·82% (52·41%-92·27%) |
| Severe COVID-19 | 0 | 3162 | 0 | 0 | 1200 | 0 | NA |

**Figure 2.**
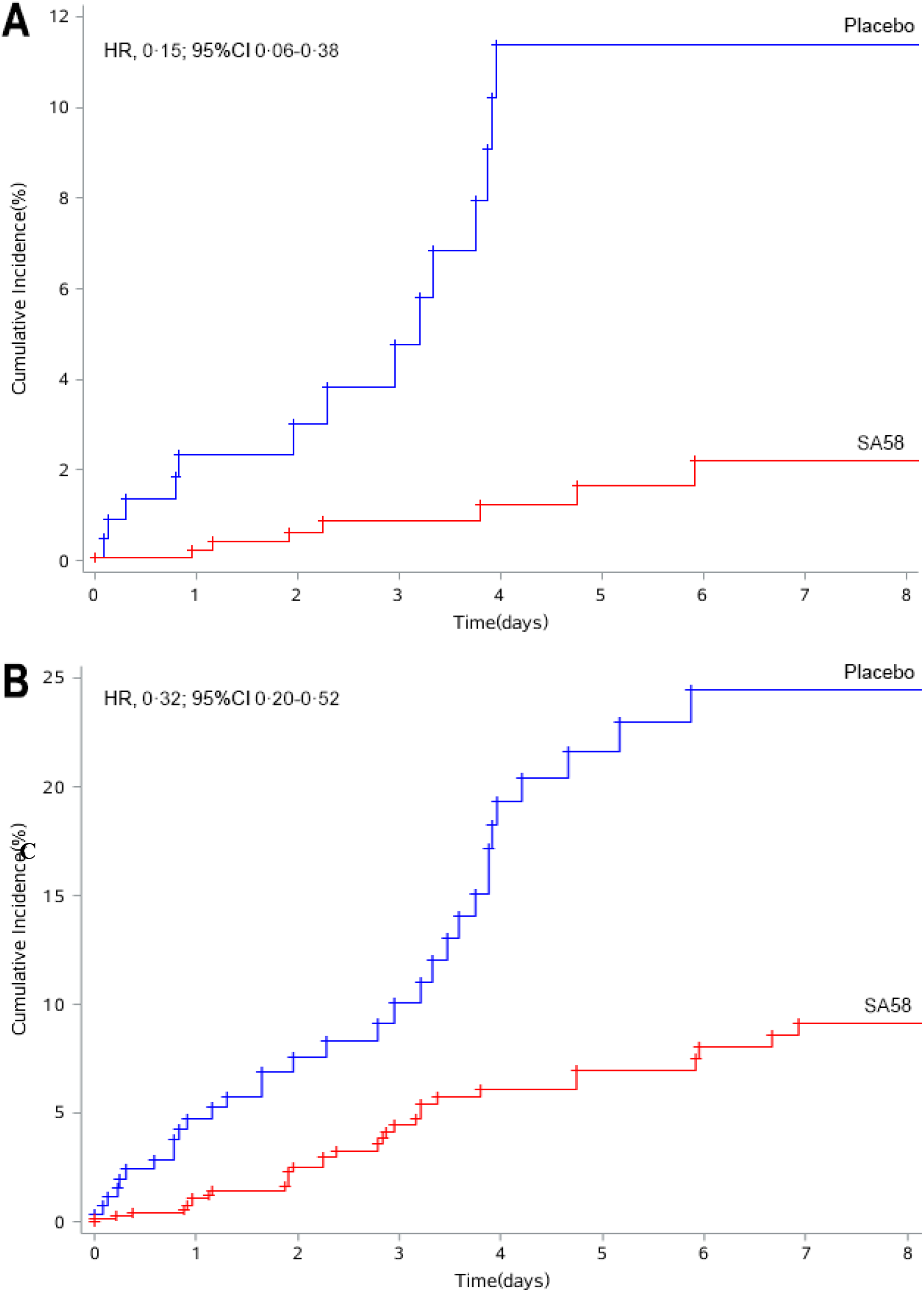
Kaplan-Meier estimates of the cumulative risk of having COVID-19. Panel A. laboratory-confirmed symptomatic COVID-19; Panel B. SARS-CoV-2 RT-PCR positive. Abbreviation, HR, Hazard ratio; 95%CI, 95% confidence interval.

No severe COVID-19 or death developed in the study participants during case monitoring period.

### Adverse Events

In the safety analysis, ≥1 AE was reported by 221 of 901 (25%) and 72 of 321 (22%) participants in the SA58 and placebo groups, respectively. All of the reported AEs were Grade one events as judged by study investigator. No SAEs and AESIs were reported by study participants, and no AEs led to study withdrawal or death during the short monitoring period. The most common AEs included nasal mucosal dryness (5%), runny nose (3%), fever (3%), nasal congestion (3%), headache (2%), cough (2%), and throat dryness (2%) etc. There was no significant difference in the occurrence of the above AEs between the SA58 and placebo groups (Table 3).

**Table 3.** Summary of Adverse Events

| Characteristics | SA58<br>(n=901) | Placebo<br>(n=321) | p-value |
| --- | --- | --- | --- |
| Treatment Emergent Adverse Events (TEAE) | 221 (25) | 72 (22) | 0·4934 |
| Nasal mucosal dryness | 50 (6) | 15 (5) | 0·6640 |
| Runny nose | 29 (3) | 11 (3) | 0·8556 |
| Headache | 26 (3) | 4 (1) | 0·1398 |
| Fever | 22 (2) | 11 (3) | 0·4216 |
| Nasal congestion | 21 (2) | 10 (3) | 0·4148 |
| Sneezing | 20 (2) | 5 (2) | 0·6465 |
| Cough | 20 (2) | 9 (3) | 0·5277 |
| Throat dryness | 20 (2) | 8 (2) | 0·8283 |
| Dizziness | 18 (2) | 5 (2) | 0·8117 |
| Sore throat | 17 (2) | 3 (1) | 0·3133 |
| SAEs | 0 | 0 | 1 |
| AESI | 0 | 0 | 1 |

Adverse events (AEs) were coded using the Medical Dictionary for Regulatory Activities, version 24·0. Abbreviations, AEs, adverse events; SAEs, severe adverse events; AESI, adverse events of special interest (i.e., allergic reaction, autoimmune reaction, and nasal and throat AEs of Grade 2 and above).

## Discussion

During our study period, the Omicron BF·7 sublineage caused large outbreaks in Beijing. Our preliminary results in healthy adult workers within 72 hours of contact with SARS-CoV-2-infected individuals showed that SA58 nasal spray was highly effective in preventing symptomatic COVID-19 and SARS-CoV-2 infection caused by Omicron BF· 7 sublineage, which variants have shown significant escape of immunity in previous studies.^17^ SA58 was able to significantly lower the risk of laboratory-confirmed COVID-19 by 80·82% (95%CI 52· 41%-92·27%) and of SARS-CoV-2 infection by 61·83% (95%CI 37·50%-76·69%) in our study participants, which has far-reaching implications. This newly developed mAb may provide a new powerful countermeasure to tackle against this cunning virus, which is currently circulating in China as a result of reopening of the country. Moreover, our study demonstrated that SA58 had a favorable safety profile and was well tolerated by healthy adults, with mild and short symptoms of nasal dryness, running nose, and nasal congestions observed among study participants. The intranasal administration of SA58 is novel and has some advantages over intramuscular injections of mAbs previously licensed, as it is less invasive and more acceptable in recipients. Auto-administration with easiness of use may allow early administration, probably a key feature for prevention.

The median incubation period of SARS-CoV-2 was estimated to be five days, with most case developing symptoms within 11·5 days of infection.^18^ The distribution of the incubation period has implications for the use of SA58 as a PEP treatment. SA58 contains mAbs that can potently neutralizes a wider range of circulating Omicron variants in vitro, including BA· 1, BA· 2, BA·2· 12·1, BA·3, BA·4/BA·5, BF·7, and other variants that have been tested so far.^14^ The mAbs is not absorbed into blood and acts as a physical barrier to stop the attachment and entry of viral particles into target cells of the throat and Nasopharyngeal mucosa. The short half-life of SA58 in nasal mucosa suggests that the effect of SA58 was transient after administration and should be administered as early as possible to cover the incubation of SARS-CoV-2 infection. Since the incubation period of the Omicron variant was reported to be as short as three days,^19^ our selection of study participants within 72 hours of contact with SARS-CoV-2-infected individual is appropriate. The duration of case monitoring is less than three days for most of study participants in the study. To evaluate the efficacy of SA58, a longer monitoring period of ≥11 days might be justified in the study.

There were 99 SARS-CoV-2 RT-PCR positive participants at baseline, suggesting that they have been infected at enrolment. These 99 participants were offered SA58 (n=77) and placebo (n=22). One purpose of PEP with mAb is to slow the illness progression prior to the illness onset. Since the sample size was low in the current analysis, we did not evaluate the effect of SA58 in lowering severity of symptoms or in shortening illness duration in these study participants. To explore the potential benefits of SA58 in slowing illness progression or shortening duration, we recommend evaluate this in upcoming studies.

Our study has several limitations. First, our study participants were confined to healthy workers generally young and healthy, which limits the generalizability of our study results to other populations, namely the elderly living in long-term care facilities, healthcare personnel who have frequent contacts with patients at increased risk of severe outcomes, and people who have underlying medical conditions. These populations have high risk of severe disease or death following SARS-CoV-2 infection and are more likely to benefit from PEP of mAbs treatment. In the future, studies need to be conducted in these high-risk populations to expand the target population of SA58. Second, concurrent practices may impact on the observed effect of SA58. For example, in an attempt to block transmission and to quickly stop outbreak, our infected study participants were managed at isolation facilities for highly infectious diseases, which lowers the risk of infection and may distort associations between treatment and disease outcome. Further assessment of SA58 in participants continuously exposed to SARS-CoV-2 in real-world situation is needed. Finally, in this preliminary analysis the sample size was low. The continuous follow-up of study participants and full-powered number of participants (e.g., n=2300) is advised to confirm the efficacy and safety of SA58.

## Conclusions

SA58 in healthy adults with early exposure to SARS-CoV-2 within 72 hours has shown satisfactory efficacy and safety in reducing symptomatic COVID-19 and SARS-CoV-2 infections. SA58 as a potential PEP treatment in preventing COVID-19 should be further evaluated in high-risk populations who are at risk of severe outcomes following infection, e.g., the elderly, healthcare personnel and people with predisposing underlying illnesses.

## Data Availability

All data produced in the present study are available upon reasonable request to the authors.

## Contributors

RS, GZ, XM, WY, and RJ designed the trial and study protocol. RS and GZ contributed to the literature search. XX, XL, ZZ, and YC verified the data. JY and XM wrote the first draft manuscript. RS, GZ, XM, XX, YC, WY, and RJ contributed to the data interpretation and revision of the manuscript. XC, ZZ, and RJ contributed to data analysis. MX monitored the trial. XM, XC, JL, and ZZ were responsible for the site work including the recruitment, follow-up, and data collection, and XL was the site coordinator. XC and ZZ were responsible for the laboratory analysis. All the authors had full access to all the data in the study and had final responsibility for the decision to submit for publication.

## Data sharing

De-identified individual participant-level data will be available upon written request to the corresponding author following publication.

## Acknowledgements

We wish to thank all the participants who took part and contribute specimens in our study.

## Reference

1. Ikematsu H, Hayden FG, Kawaguchi K, et al. Baloxavir Marboxil for Prophylaxis against Influenza in Household Contacts. N Engl J Med 2020; 2020(4): 309–20.

2. World Health Organization. WHO Expert Consultation on Rabies: third report. World Health Organization; 2018. https://apps.who.int/iris/bitstream/handle/10665/272364/9789241210218-eng.pdf

3. Kuhar DT, Henderson DK, Struble KA, et al. Updated U.S. Public Health Service guidelines for the management of occupational exposures to HIV and recommendations for postexposure prophylaxis.2018. https://stacks.cdc.gov/view/cdc/20711

4. Redeker AG, Mosley JW, Gocke DJ, McKee AP, Pollack W. Hepatitis B immune globulin as a prophylactic measure for spouses exposed to acute type B hepatitis. N Engl J Med 1975; 1975(21): 1055–9.

5. Brunell PA, Ross A, Miller LH, Kuo B. Prevention of varicella by zoster immune globulin. N Engl J Med 1969; 1969(22): 1191–4.

6. Slifka MK, Amanna IJ. Passive Immunization. In: Orenstein W, Offit PA, Edwards KM, Plotkin SA, eds. Plotkin’s Vaccines. 7th ed: Elsevier Health Sciences; 2018: 84–95.

7. Zhu N, Zhang D, Wang W, et al. A Novel Coronavirus from Patients with Pneumonia in China, 2019. The New England journal of medicine 2020; 2020(8): 727–33.

8. Walker PGT, Whittaker C, Watson OJ, et al. The impact of COVID-19 and strategies for mitigation and suppression in low- and middle-income countries. Science 2020; 2020(6502): 413–22.

9. World Health Organization. WHO Coronavirus (COVID-19) Dashboard. 2022. https://covid19.who.int/.

10. Strohl WR, Ku Z, An Z, Carroll SF, Keyt BA, Strohl LM. Passive Immunotherapy Against SARS-CoV-2: From Plasma-Based Therapy to Single Potent Antibodies in the Race to Stay Ahead of the Variants. BioDrugs 2022; 2022(3): 231–323.

11. U.S. Food and Drug Administration. Coronavirus (COVID-19) | Drugs. 2022. https://www.fda.gov/drugs/emergency-preparedness-drugs/coronavirus-covid-19-drugs

12. U.S. Food and Drug Administration. Emergency Use Authorization. 2022. https://www.fda.gov/emergency-preparedness-and-response/mcm-legal-regulatory-and-policy-framework/emergency-use-authorization#coviddrugs

13. Levin MJ, Ustianowski A, Thomas S, et al. AZD7442 (Tixagevimab/Cilgavimab) for Post-exposure Prophylaxis of Symptomatic COVID-19. Clinical infectious diseases. 2022.

14. Cao Y, Jian F, Zhang Z, et al. Rational identification of potent and broad sarbecovirus-neutralizing antibody cocktails from SARS convalescents. Cell Rep 2022: 111845.

15. National Health Commission of the People’s Republic of China. Protocol for Prevention and Control of COVID-19 (Edition 9). Beijing: National Health Commission of the People’s Republic of China; 2022. http://www.nhc.gov.cn/yzygj/s7653p/202203/b74ade1ba4494583805a3d2e40093d88.shtml.

16. NIH National Cancer Institute. Common Terminology Criteria for Adverse Events (CTCAE) Version 5.0. 2017. https://evs.nci.nih.gov/ftp1/CTCAE/CTCAE_5.0.

17. Cao Y, Wang J, Jian F, et al. Omicron escapes the majority of existing SARS-CoV-2 neutralizing antibodies. Nature 2022; 2022(7898): 657–63.

18. Lauer SA, Grantz KH, Bi Q, et al. The Incubation Period of Coronavirus Disease 2019 (COVID-19) From Publicly Reported Confirmed Cases: Estimation and Application. Ann Intern Med 2020; 2020(9): 577–82.

19. Xin H, Wang Z, Feng S, et al. Transmission dynamics of SARS-CoV-2 Omicron variant infections in Hangzhou, Zhejiang, China, January-February 2022. International journal of infectious diseases : IJID 2022; 126: 132–5.

