## Supplementary material for "Post-Exposure Prophylaxis with SA58 (anti-COVID-19 monoclonal antibody) Nasal Spray for the prevention of symptomatic Coronavirus Disease 2019 in healthy adult workers: A randomized, single-blind, placebo-controlled clinical study"

This appendix has been provided by the authors to give readers additional information about their work.

### Supplementary methods

#### Full inclusion criteria

A participant had to meet the following criteria to be eligible for inclusion in the study:

- Age ≥18 years at the time of enrolment.
- Potential exposure to a specific identified individual with laboratory-confirmed SARS-CoV-2 infection (index case), and the presumably contact occurred within 72 hours of the positive RT-PCR test of the index case.
- Prior to enrollment, participants must not have had COVID-19 symptoms, as described in **Supplementary Table 1**.
- Participants must be voluntary to participate in the study, and provide written informed consents before enrolment.

#### Full exclusion criteria

A participant who met any of the following criteria was excluded from the study:

- individuals with known history of severe allergies or reaction to any component of inhaled SA58 nasal spray.
- those currently pregnant, lactating or expected to be pregnant during the study period.
- those who participated in any kind of clinical trials of SARS-CoV-2 neutralizing antibody injections in the preceding 180 days before screening or participated in any investigational medicinal product in the preceding four weeks before screening.
- were unable to take nasal spray inhalation.
- had reported fever at enrolment or axillary temperature of more than 37·0 °C.
- had severe neurological disease (e.g., epilepsy, convulsions, or seizures) or psychosis, or family history of psychosis.
- or had any other significant chronic disease, disorder, or finding that, in the judgment of the investigator, significantly increased the risk to the participant because of participation in the study, affected the ability of the participant to participate in the study, or impaired interpretation of the study data.

#### Laboratory methods for sequencing of SARS-CoV-2 variants

Genomic RNA of positive RT-PCR sample was isolated by QIAamp Viral RNA Mini Kit (250) (QIAGEN), then reverse transcripted to ds cDNA using ULSEN® Ultra Sensitive Whole Genome Capture Kit for SARS-CoV-2 (MicroFuture). Genome cDNAs were digested, added sequencing primers and sequenced following the instruction of Illumina NextSeqTM 550 and Illumina Kits. Genomic sequencing data were combined and assembled using software of MicronCoV® Analyzer (MicroFuture), then identified the mutant clades using Nextclade online database (https://clades·nextstrain·org/).^1^

### Supplementary tables

#### Supplementary Table 1. Symptoms of COVID-19. Participant must present with ≥ two of the type A symptoms, or at least one of the type B symptoms, or with chest imaging characteristics suggestive of COVID-19, and with a positive RT-PCR test of SARS-CoV-2.

| Type A symptoms (no minimum duration) | Type B symptoms (must be present for ≥ two days) |
| --- | --- |
| - Fever - Chills - Sore throat - Fatigue - Runny nose or nasal congestion - Muscle aches - Headache - Nausea or Vomiting - Diarrhea | - Cough - New loss of smell - New loss of taste - Shortness of breath (no minimum duration required) - Difficulty breathing (no minimum duration required) |
| Chest imaging characteristics of COVID-19. In the early stage, there were multiple small patches and interstitial changes, especially in the lung periphery. Furthermore, it develops into multiple ground glass shadows and infiltrating shadows in both lungs. In severe cases, lung consolidation may occur, and rarely pleural effusion. In multisystem inflammatory syndrome in children (MIS-C), heart shadow enlargement and pulmonary edema can be observed in patients with cardiac insufficiency. | |

Abbreviations: COVID-19, coronavirus disease 2019. RT-PCR, reverse-transcription polymerase chain reaction. SARS-CoV-2, severe acute respiratory syndrome coronavirus 2.

#### Supplementary Table 2. Severe COVID-19 defined according to the Protocol for Prevention and Control of COVID-19 (9^th^ edition)

| **Patient state** | **Description** |
| --- | --- |
| Mild case | The clinical symptoms were mild, and there was no evidence of pneumonia on chest imaging. |
| Moderate case | The clinical symptoms were mild, and there was evidence of pneumonia on chest imaging. |
| Severe case | Shortness of breath, respiratory rate ≥30 times/min; |
|  | In the resting state, when inhaling air, the oxygen saturation is ≤93%. |
|  | The ratio of arterial oxygen partial pressure (PaO_2_) to fractional inspired oxygen (FiO_2_) is ≤300mmHg (1mmHg=0·133kPa). |
|  | Progressive of clinical symptoms, with lung imaging shows that the lesion progresses >50% within 24-48 hours. |
| Very-severe case | Respiratory failure occurs and mechanical ventilation is required. |
|  | Shock |
|  | Other organ failure requires ICU monitoring and treatment |

Severe COVID-19 was defined as severe case and very-severe case combined. Abbreviations, FiO_2_, fraction of inspired oxygen; PaO_2_, partial pressure of oxygen; ICU, intensive care unit.

#### Supplementary figure 1 Phylogenetic tree of SARS-CoV-2 strains identified in the study produced by using Nextclade.

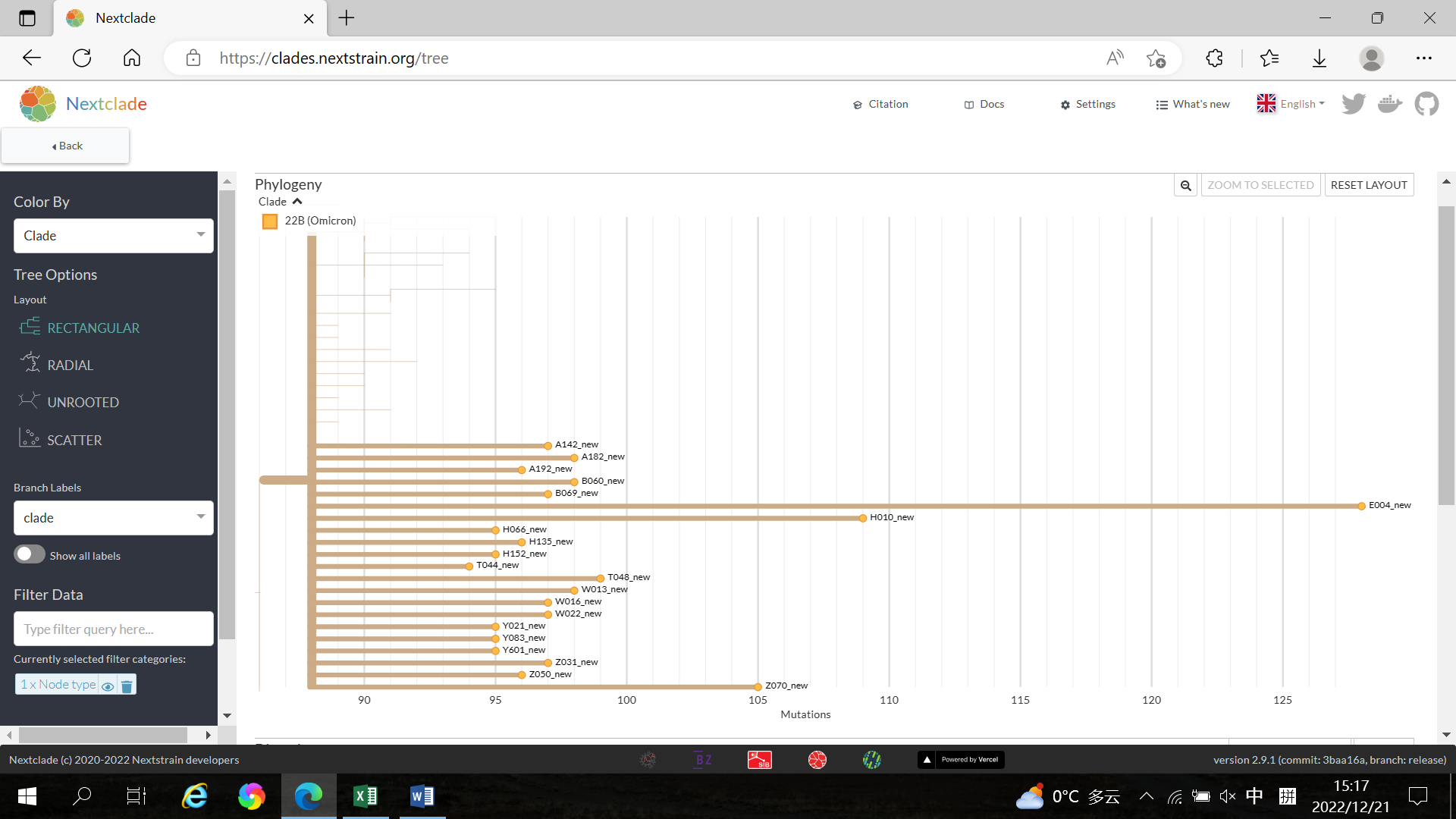

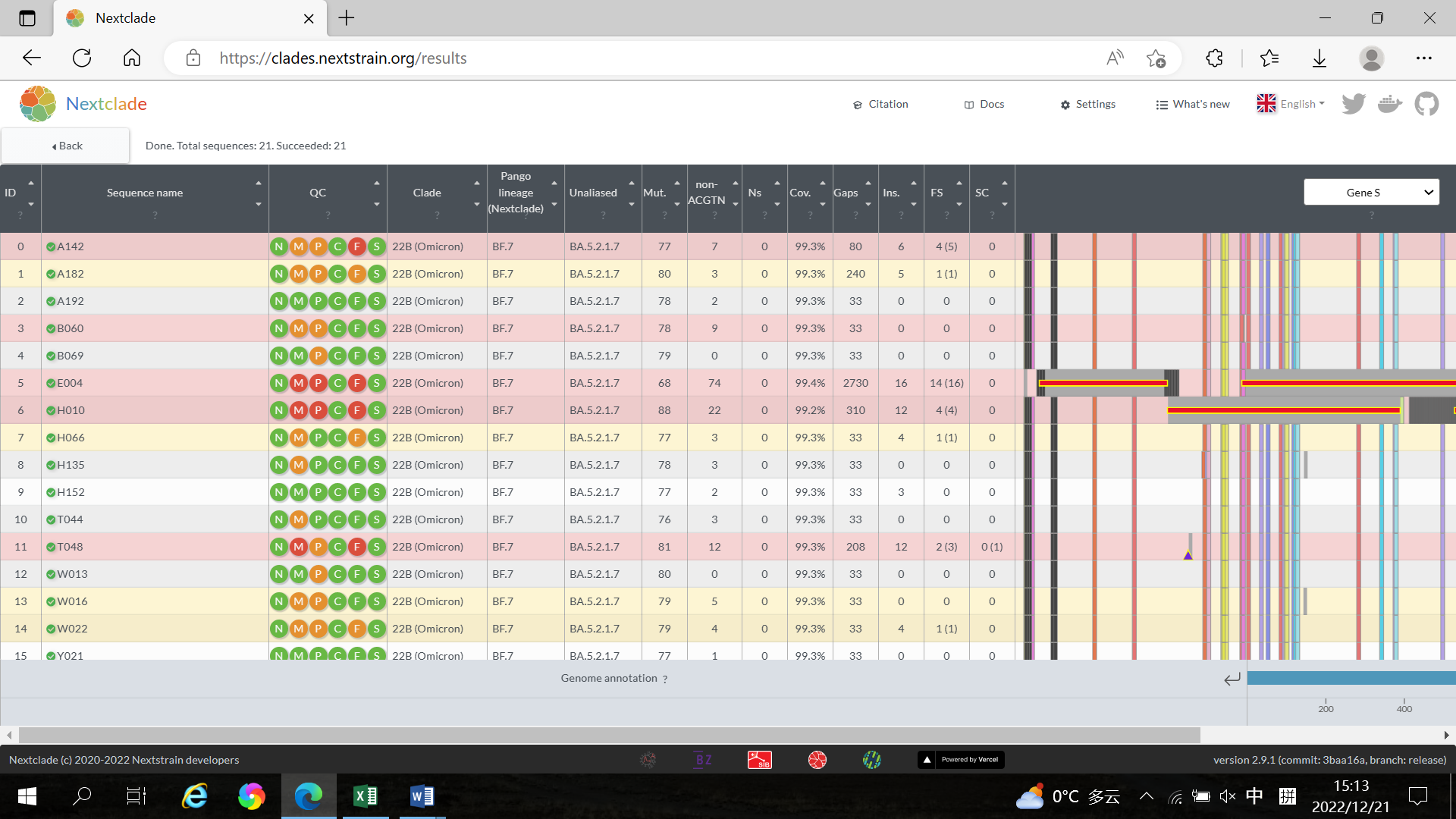

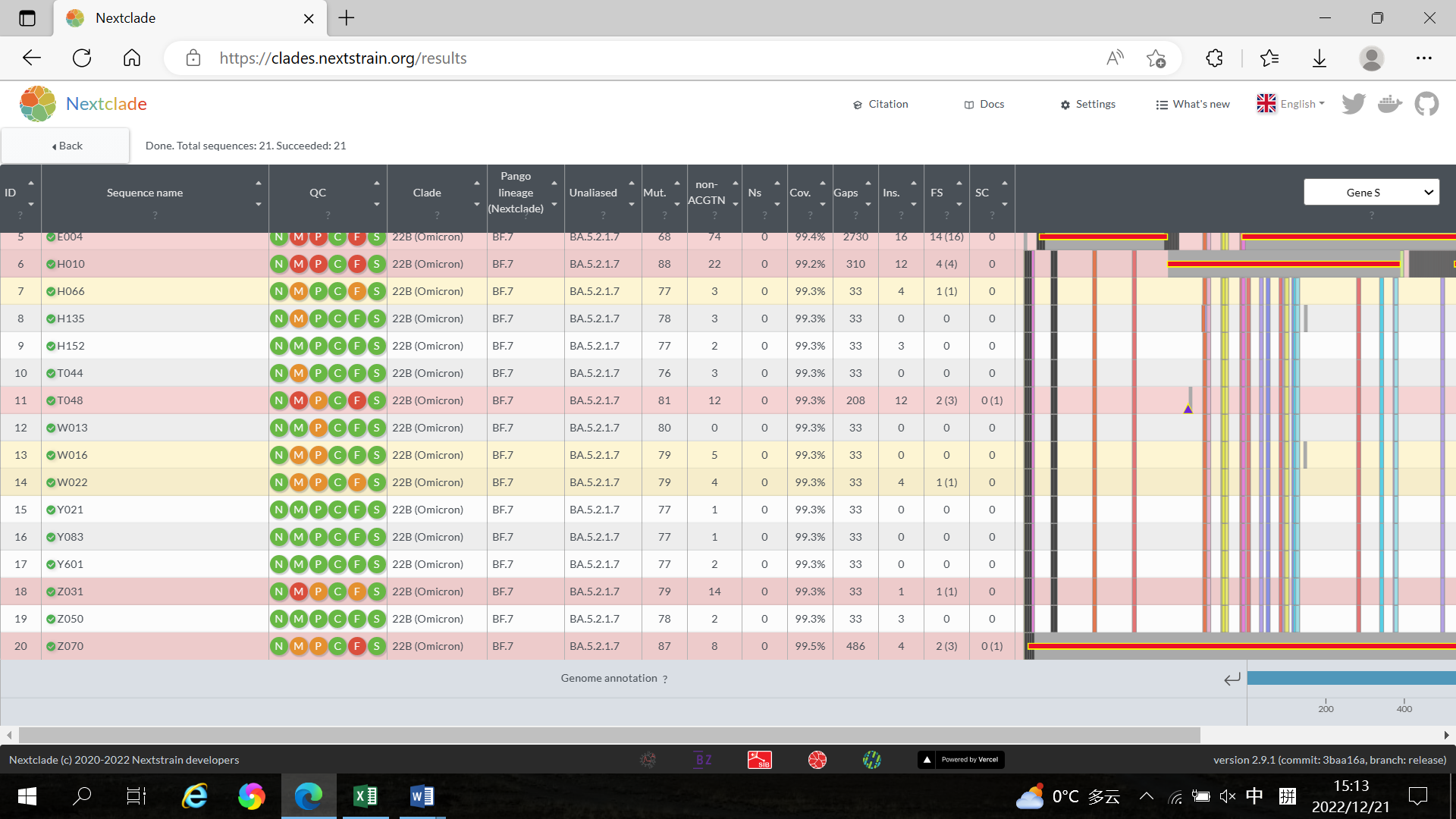

Green: good, Orange: mediocre, Red: bad. N：missing data；M：mixed sites；F：frame shifts；P：private mutations；C：mutation clusters；S：stop codons.

### Reference

1. Hadfield J, Megill C, Bell SM, et al. Nextstrain: real-time tracking of pathogen evolution. *Bioinformatics* 2018; **34**(23): 4121-3. doi: 10.1093/bioinformatics/bty407.
